# Modelling brain stimulation in cerebral palsy: electric field insights from paediatric tDCS

**DOI:** 10.64898/2026.08.07.26359953

**Authors:** Matthew Weightman, Bronwyn Gavine, Foteini Mavrommati, Heidi Johansen-Berg, Helen Dawes, Melanie K Fleming

## Abstract

**Background:** Transcranial direct current stimulation (tDCS) is increasingly used as an adjunct to rehabilitation for young people with cerebral palsy (CP), yet considerable variability exists in clinical response. Individualised electric field modelling provides an opportunity to estimate the distribution of electrical fields generated by the stimulation delivered to the brain and explore potential relationships with functional outcomes.

**Methods:** Structural MRI scans from nineteen participants (10-16 years) from a previously published randomised controlled trial (ISRCTN74235136) investigating the effects of tDCS combined with motor training, underwent participant-specific finite element modelling using SimNIBS. Electric field strength was quantified within anatomically defined motor regions of interest, including the primary motor cortex (M1), dorsal premotor cortex (PMd), supplementary motor area (SMA), and a combined motor network. Global grey matter electric field metrics and stimulation focality were also extracted.

**Results:** Estimated electric field strength differed significantly across motor regions (p<0.001), with PMd receiving significantly greater stimulation than both M1 and SMA. Electric field strength within a control region (primary visual cortex) was significantly lower than within M1 (p<0.001). Despite inter-individual variability in regional and global electric field metrics, no significant associations were observed between estimated electric field strength or focality and changes in function following intervention.

**Conclusion:** Individualised electric field modelling demonstrated that an M1-targeted tDCS montage preferentially stimulated PMd rather than M1 in young people with CP. These findings highlight the importance of subject-specific modelling when characterising current distribution and suggest that variability in electric field strength alone does not explain variability in behavioural response.

**Highlights:**

- Individualised modelling characterised tDCS electric fields in paediatric CP.
- M1-targeted tDCS produced greater electric field strength in PMd than M1.
- Motor regions received greater electric field strength than control visual cortex.
- Electric field strength did not predict upper- or lower-limb functional change.

## Introduction

Cerebral palsy (CP) is a heterogeneous, lifelong, neurodevelopmental disorder characterised by impairments of movement and posture resulting from non-progressive injury to the developing brain [1]. Clinical manifestations may include spasticity, dystonia, choreoathetosis, ataxia, or a combination of these motor phenotypes [2]. In addition to motor impairments, individuals with CP often experience associated sensory, cognitive, and affective difficulties, which further impacts their functional independence and quality of life [3]. Despite efforts to prevent the incidence of CP, it remains the most common cause of physical disability in childhood and represents a major contributor to paediatric morbidity worldwide [4, 5]. Although there have been advances in rehabilitation, improvements in function are often modest and highly variable between individuals [6, 7]. Given this, it is essential to develop adjuvant interventions to enhance motor function and subsequently independence and quality of life of young people with CP.

Transcranial Direct Current Stimulation (tDCS) is a form of non-invasive brain stimulation used to modulate ongoing neural excitability [8, 9]. tDCS is safe, painless, and relatively inexpensive. Accordingly, there has been great interest in its therapeutic application, with evidence of its potential efficacy across a spectrum of neurological disease/disorders [10–12]. With respect to CP, tDCS appears to be safe, with guidelines developed for its implementation [13, 14]. However, robust research on the effectiveness of tDCS on motor function is limited, with mixed findings and restricted to specific CP subtypes [15–17]. Understanding this inconsistency is therefore critical to optimising protocols and maximising therapeutic potential.

The inconsistent effects of tDCS may, at least in part, reflect the between-subject variability in the electrical dose delivered to the intended cortical target. Most tDCS studies in CP aim to modulate the excitability within the primary motor cortex (M1), typically contralateral to the most affected side of the body [15]. However, current flow through the brain is strongly influenced by individual anatomy, including skull thickness, cortical folding, lesion characteristics, cerebrospinal fluid distribution, and brain volume [18–20]. Consequently, identical electrode montages applied across individuals may produce markedly different electric field distributions within the brain. Without computational modelling, it is difficult to determine whether stimulation reaches the intended cortical regions or whether inter-individual differences in electric field delivery contribute to the variability in behavioural response.

MRI-based finite element modelling enables subject-specific estimation of electric field magnitude and spatial distribution throughout the brain [21]. Previous work has demonstrated substantial variability in electric field strength between individuals and has highlighted discrepancies concerning intended and actual stimulation targets [20, 22, 23]. Such approaches have been used increasingly to characterise dose delivery, evaluate stimulation focality, and investigate dose-response relationships between electric field strength and behavioural outcomes. However, relatively few studies have estimated individualised electric field modelling in paediatric CP populations, where neuroanatomical heterogeneity and lesion-related grey/white matter alterations may be especially important. Developmental research has suggested that peak electric field strength and focality may vary between adults and children, likely due to differences in skull thickness and other age-related anatomical changes [24]. To our knowledge, only two studies have investigated tDCS-induced electric fields in CP [14, 25], providing practical information regarding optimal montage selection in paediatric brains with differing pathologies. However, evaluation of the technique is limited, and specifically whether inter-individual variability in modelled electric field strength within targeted motor regions is associated with responsiveness to intervention remains unknown.

Therefore, the aim of the present study was to use individualised MRI-based finite element modelling to characterise estimated electric field strength and focality in a cohort of young people with CP receiving anodal tDCS targeting M1. These data are from a published clinical trial investigating the effects of tDCS with upper and lower limb motor training on motor function [26]. We quantified electric field strength within anatomically defined motor regions of interest and compared estimated stimulation magnitude across M1, premotor cortex (PMd), and supplementary motor area (SMA). We additionally examined whether inter-individual variability in modelled electric field strength within ROIs was associated with changes in functional motor performance improvements.

We hypothesised that our montage would generate stronger estimated mean electric fields within M1 compared to neighbouring motor regions, and a greater strength of tDCS-induced electric field within the targeted M1 would correlate with more improvement in upper and lower limb function following 7-10 sessions of tDCS and motor training.

## Methods

### Study Design

Data used in the present analyses were derived from a previously published two-arm, parallel-group, randomised controlled pilot trial investigating the effects of transcranial direct current stimulation (tDCS) combined with motor training in young people with cerebral palsy [26]. The study was designed and reported in accordance with CONSORT guidelines, approved by the United Kingdom National Research Ethics Service (West Midlands - Edgbaston Research Ethics Committee; reference 20/WM/0046), and prospectively registered on the ISRCTN clinical trials registry (ISRCTN74235136) prior to enrolment of the first participant. The present study represents a secondary analysis of MRI-derived electric field modelling data obtained from participants enrolled in the trial. In short, participants aged 10-16 years with cerebral palsy affecting the upper and/or lower limbs (n = 27) were randomised to receive either active (n = 14) or sham (n = 13) transcranial direct current stimulation (tDCS). Stimulation was delivered over 10 sessions in conjunction with upper- and lower-limb motor training. Functional outcomes were assessed at baseline and one week following completion of the intervention. The primary clinical outcomes were upper-limb function, measured using the Jebsen-Taylor Hand Function Test (JTT; [27]), and functional mobility, measured using the Timed Up and Go (TUG; [28]) test. Full details regarding participant recruitment, eligibility criteria, randomisation procedures, intervention protocols, and primary clinical outcomes have been reported previously [26].

### Participant Demographics

20 of the 27 participants in the original trial opted to be involved in the MRI component of the study. Of these 20 participants, 19 met the inclusion criteria of having useable T1 and T2-weighted structural MRI scans and completed outcomes at the 1-week follow up assessment (7 females; mean age ± standard deviation: 12.6 ± 1.6 years). Of the 19 participants 11 were randomly allocated into the active (anodal) stimulation group (4 females; mean age ± standard deviation: 13.1 ± 1.6 years), and 8 the sham group (3 females; mean age ± standard deviation: 11.9 ± 1.4 years). At baseline the average Gross Motor Function Classification System (GMFCS) level was 1.95 (standard deviation ± 0.52; range = 1-3) and Manual Ability Classification System (MACS) score was 1.76 (standard deviation = ± 0.75; range = 1-3), representing a mild to moderate level of functional impairment across the cohort.

### Electric Field Modelling

#### Imaging

Magnetic resonance imaging (MRI) was conducted at the University of Oxford Centre for Integrative Neuroimaging. MRI data were acquired in a single scanning session prior to the start of the study intervention. Data were collected using a 32-channel head coil with 3.0-T Prisma Magnetom Siemens scanners, software version VE11C (Siemens Medical Systems, Erlangen, Germany). Head movement was minimised using foam padding and limb strapping as required. The T1w structural scan sequence was acquired with a Magnetization Prepared Rapid Acquisition Gradient Echo (MPRAGE) sequence (TR = 1900ms, TE = 3.97ms, voxel size = 1.0 x 1.0 x 1.0mm, flip angle = 8°, total slices = 192, field of view (FOV) = 192mm^3^). T2-weighted images were acquired in the sagittal plane (TR = 5000ms; TE = 397ms; voxel size = 1.0 x 1.0 x 1.0mm; slice thickness = 1.05mm; 192 slices; FOV = 256mm^3^). Diffusion and resting state scans were also acquired but not required for the present analyses.

#### Head Model Generation

Individualised head models were generated from each participant’s T1- and T2-weighted structural MRI scans using SimNIBS (version 4.6.0; [29–31]) and the CHARM pipeline [32]. CHARM combines atlas-guided segmentation with surface reconstruction to produce subject-specific finite element head models, including representations of scalp, skull, cerebrospinal fluid, grey matter, and white matter. The resulting segmentations were converted into tetrahedral meshes for subsequent electric field simulation (Figure 1). All segmentation results and head models were visually inspected to confirm anatomical plausibility and quality prior to further analysis. Structural lesions were identified in two participants, and lesion masks were manually delineated in native T1 space prior to mesh generation. These masks were incorporated into the segmentation workflow to improve tissue classification within the resultant head models.

**Figure 1:**
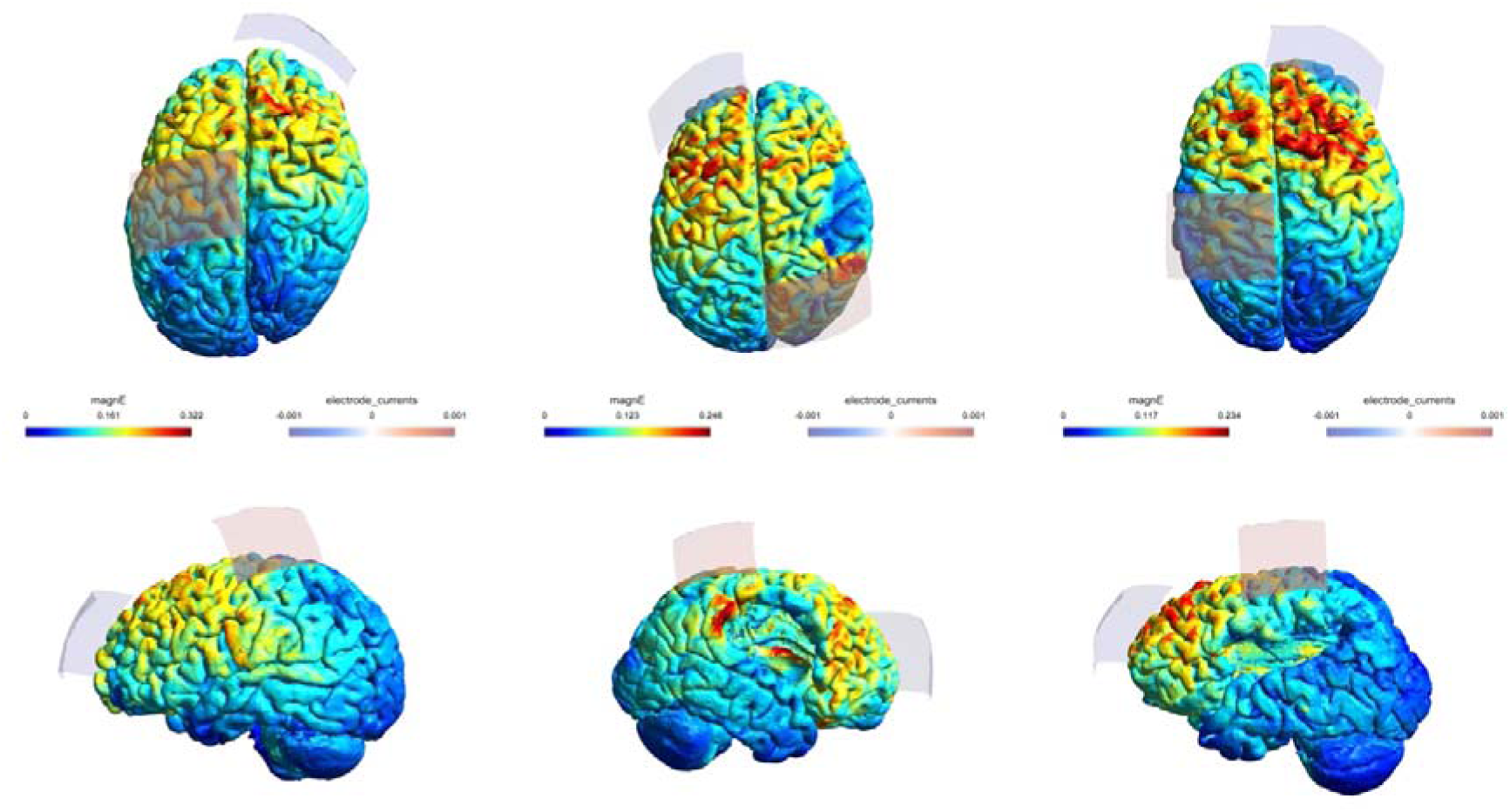
Example electric field magnitude (V/m) distributions projected onto the cortical surface (lateral and superior views) of 3 participant with differing pathologies following finite element modelling (SimNIBS: CHARM pipeline). Grey shapes represent electrode locations. The anode was positioned over the primary motor cortex contralateral to the more-affected side (C1/C2), with the cathode positioned over the contralateral supraorbital region (AF3/AF4). Note the anode location of CP2 was used in one subject (middle) to avoid direct placement over lesioned cortex.

#### Electrode Montage and Simulation Parameters

Participants allocated to the active intervention group received anodal tDCS (1 mA; Neurostym, Brainbox Ltd, UK) during the first 20 minutes of each 90 min motor training session. The stimulation montage was designed to target the primary motor cortex contralateral to the participant’s more-affected side, with the intention of influencing both upper- and lower-limb motor representations. The anode (5 x 7cm; 35cm²) was centred over C1 or C2, as determined using the international 10-20 EEG system, and positioned as close to the sagittal midline as possible. The cathode (5 x 7 cm; 35 cm²) was positioned over the contralateral supraorbital region, corresponding approximately to AF3 or AF4 in our paediatric cohort. For one participant, a large lesion adjacent to the intended motor target was identified. Consequently, the anode position was adjusted 1cm posteriorly (CP2) to avoid direct placement over the lesion while maintaining stimulation of the ipsilesional motor network.

Participants allocated to the sham condition received electrode placement identical to the active condition, however, the stimulation current was ramped up over 30 seconds and then turned off, thereby mimicking the initial sensation of stimulation without delivering a sustained current. Electrodes remained in place for the same duration as the active condition.

Modelling: Electrode montages were modelled using a multilayer array consisting of a 1.5mm rubber electrode positioned between two 4mm saline-soaked (0.9% NaCl) sponge layers. Standard SimNIBS conductivity values were used [20, 33, 34]. Following simulation, electric field distributions and electrode placements were visually inspected to confirm appropriate positioning and current flow patterns. Finite element simulations were performed using SimNIBS to generate participant-specific estimates of electric field magnitude throughout the brain (Figure 1). Electric field magnitude was used as the primary modelling outcome measure.

#### ROI Definition and Electric Field Extraction

Cortical regions-of-interest (ROIs) were identified using the Human Connectome Project Multi-Modal Parcellation atlas (HCP-MMP1; [35]), projected into individual participant space. Three motor ROIs were defined a priori: primary motor cortex (M1), dorsal premotor cortex (PMd), and supplementary motor area (SMA) for the targeted hemisphere. These regions were selected because they comprise key cortical nodes within the motor network responsible for the planning, preparation, and execution of voluntary movement [36]. Furthermore, their close anatomical proximity to the intended stimulation target enabled characterisation of electric field distribution across the broader motor network, allowing assessment of both target specificity and current spread beyond M1. A combined motor network ROI (Motor All) was additionally created by combining all motor parcels. A lateralised primary visual cortex (V1) ROI was also defined as a control region within the same hemisphere as the stimulated motor regions. ROI definitions were based on HCP-MMP1 parcels and are provided in Table 1.

**Table 1.** HCP-MMP1 ROI Parcels for targeted hemisphere.

| Region of Interest (ROI) | HCP-MMP1 parcels |
| --- | --- |
| Primary Motor Cortex (M1) | 4, 55b, 6d, 6v |
| Dorsal Premotor Cortex (PMd) | 6a, i6-8, s6-8, 8Av, 8C |
| Supplementary Motor Area (SMA) | 6ma, 6mp |
| Motor All | Combination of above |
| Primary Visual Cortex (V1) | V1, V1d, V1v |

Atlas registration and ROI localisation were visually inspected in all participants by projecting atlas-derived ROIs onto the individual cortical surfaces and verifying anatomical correspondence with the underlying anatomy. Electric field magnitude values (magnE; V/m) were extracted from the cortical surface mesh generated by SimNIBS. For each ROI, all surface nodes belonging to the ROI were identified and the corresponding magnE values extracted. Area-weighted mean electric field strength was calculated using the surface area associated with each node, thereby accounting for differences in mesh density across the cortical surface. In addition, the 95th percentile and maximum magnE values were calculated for each ROI to characterise higher-intensity stimulation within the region.

#### Global Electric Field Metrics

Global grey matter electric field metrics were extracted using the SimNIBS field summary framework. These included the 95th, 99th, and 99.9th percentile grey magnE values and measures of stimulation focality, defined as the volume of grey matter exposed to electric field strengths ≥50% and ≥75% of the 99.9th percentile field.

### Statistical Analyses

All statistical analyses were performed using Python (3.9.19) and R (version 4.6.0). Descriptive statistics are presented as mean ± standard deviation (SD) unless otherwise stated. Statistical significance was set at p < 0.05.

To compare electric field strength across motor regions, linear mixed-effects models were fitted using the ‘lme4’ package [37]. Prior to linear mixed-effects modelling, distributions of ROI electric field magnitude values were assessed using Shapiro-Wilk tests and were deemed normally distributed (all p > 0.05). Electric field magnitude was entered as the dependent variable, ROI (M1, PMd, and SMA) as a fixed effect, and participant as a random intercept to account for repeated measurements within individuals. A second model additionally included stimulation side and lesion status (a binary variable, defined as identification of a visible structural) as fixed effects. Omnibus effects were assessed using the ‘emmeans’ package, with Tukey-adjusted pairwise comparisons to compare electric field strength between ROIs. As a control analysis, electric field strength within the primary visual cortex (V1) was compared with M1 using a paired-samples t-test to assess the anatomical specificity of the stimulation montage.

Relationships between modelled electric field metrics and behavioural outcomes were examined within the active stimulation group only (n = 11). Behavioural change scores (%) were calculated by subtracting the one-week follow up performance from the baseline for the JTT and TUG test, with positive values therefore indicating functional improvement (quicker time to complete at follow up). Given the small sample, associations between electric field metrics and behavioural outcomes were assessed using Spearman’s rank correlation coefficients. Correlations between M1 electric field strength and behavioural change were specified a priori as the primary analyses. Associations involving PMd, the combined motor network ROI (Motor All), global grey matter electric field strength, and focality metrics were considered secondary exploratory analyses.

## Results

### Region of Interest Analyses

Estimated electric field strength differed significantly across motor ROIs (Figure 2). Mean electric field magnitude was 0.130 ± 0.027 V/m within M1, 0.148 ± 0.029 V/m within PMd, and 0.133 ± 0.024 V/m within SMA. A linear mixed-effects model demonstrated a significant main effect of ROI on electric field magnitude (F(2,40.1) = 42.37, p < 0.001). Post hoc Tukey-adjusted comparisons revealed significantly greater electric field strength within PMd than both M1 (mean difference = 0.018 V/m, p < 0.001) and SMA (mean difference = 0.015 V/m, p < 0.001). No significant difference was observed between M1 and SMA (mean difference = 0.003 V/m, p = 0.286).

**Figure 2:**
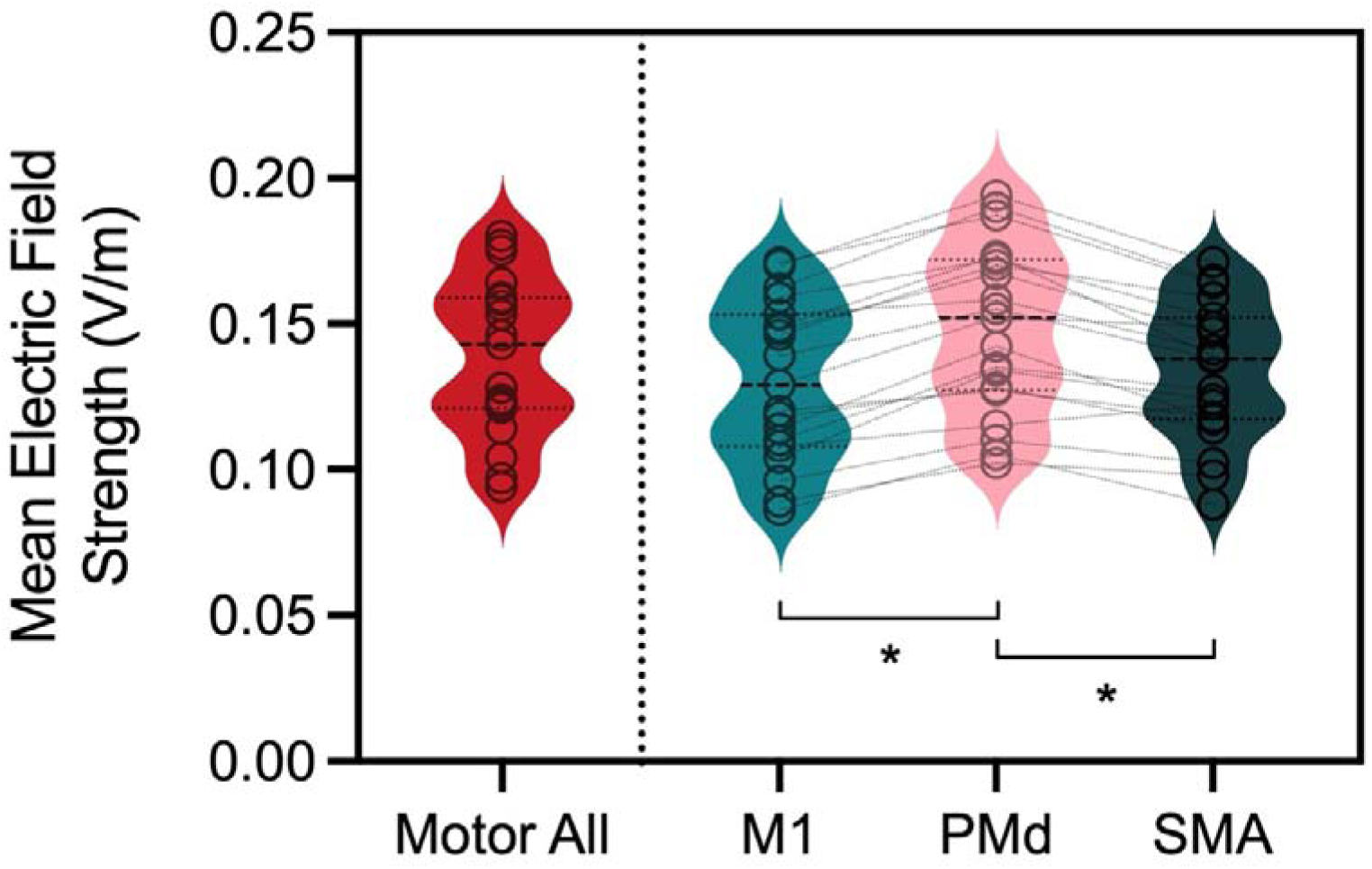
Violin plots showing the distribution of mean electric field magnitude (V/m)) across participants within the combined motor network ROI (Motor All), primary motor cortex (M1), dorsal premotor cortex (PMd), and supplementary motor area (SMA). Individual participant values are shown as circles and connected by lines across motor ROIs. Electric field strength differed significantly across motor regions, with PMd demonstrating significantly greater electric field magnitude than both M1 and SMA (*p < 0.001). No significant difference was observed between M1 and SMA. Thick dashed and thin dotted horizontal lines indicate group medians and quartiles respectively.

Inclusion of stimulation side and lesion status did not significantly improve overall model fit (χ²(2) = 3.85, p = 0.146). Stimulation side was not associated with electric field strength (p = 0.81), whereas those with visible cortical lesions demonstrated lower electric field magnitudes, although this did not prove significant (p = 0.071) and may reflect the low number of participants with lesions in our cohort (n = 2).

Electric field strength within V1 was significantly lower than M1 (paired t-test: t(18) = 12.54, p < 0.001), confirming preferential stimulation of the motor network.

### Global Electric Field Characteristics

Considerable inter-individual variability was observed in both grey matter electric field strength and focality across the cohort. The mean 99.9th percentile grey matter electric field magnitude was 0.274 ± 0.042 V/m, ranging from 0.206 to 0.337 V/m. The volume of grey matter exposed to electric field strengths equal to or exceeding 75% of the 99.9th percentile field ranged from 6.55 to 18.20cm³ (mean = 12.75 ± 2.93cm³), indicating variability in stimulation focality between participants (Figure 3).

**Figure 3:**
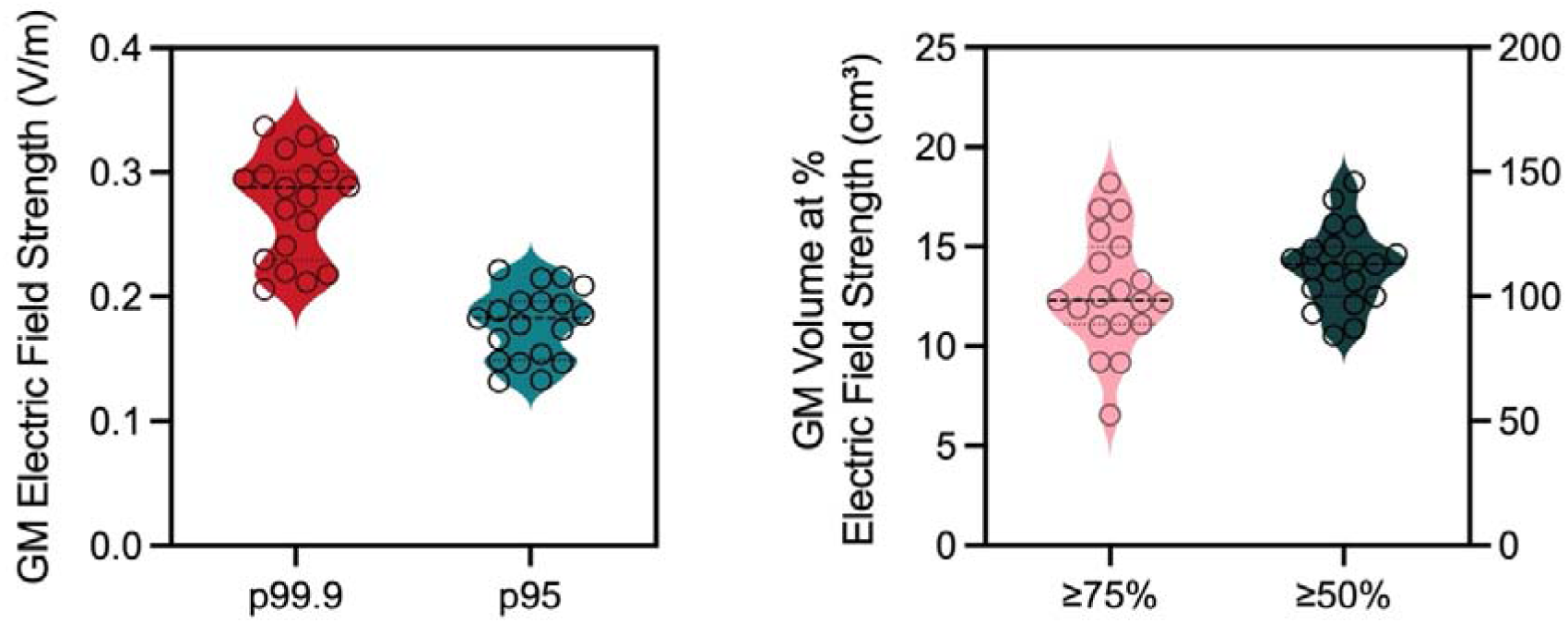
Distribution of global grey matter electric field metrics across participants. Left panel: 99.9th and 95th percentile grey matter electric field magnitude (V/m). Right panel: stimulation focality (cm^3^), defined as the volume of grey matter exposed to electric field strengths equal to or exceeding 75% and 50% of the 99.9th percentile electric field. Individual participant values are shown as circles and violin plots illustrate the distribution of values across the cohort. Thick dashed and thin dotted horizontal lines indicate group medians and quartiles respectively.

### Relationships between Electric Field Strength and Behavioural Outcomes

#### ROIs

To determine whether electric field strength within the targeted M1 was associated with functional improvements following intervention, Spearman’s rank correlations were performed between mean M1 electric field magnitude and changes in behavioural outcomes (Figure 4). No significant relationships were observed between M1 electric field strength and change in JTT total time (rho = 0.118, p = 0.735) or TUG total time (rho = -0.027, p = 0.946). Similarly, no significant associations were observed between behavioural change and electric field strength within PMd (JTT: rho = -0.100, p = 0.776; TUG: rho = -0.073, p = 0.838) or the combined motor network ROI (Motor All; JTT: rho = -0.064, p = 0.860; TUG: rho = - 0.146, p = 0.673).

**Figure 4:**
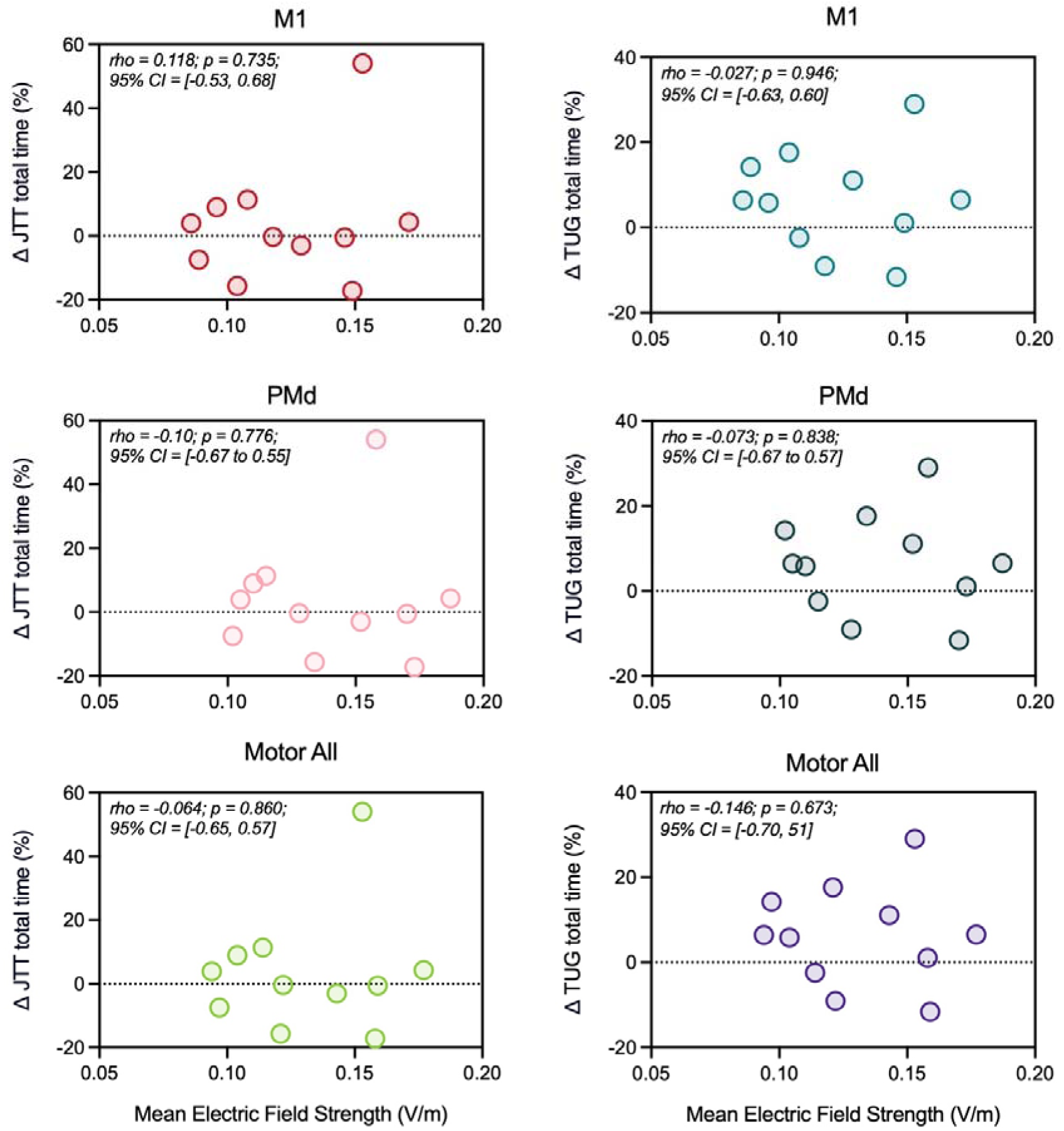
Scatterplots illustrating the relationships between mean electric field magnitude (V/m) within the primary motor cortex (M1), dorsal premotor cortex (PMd), and combined motor network ROI (Motor All) and changes in upper-limb function (Jebsen–Taylor Hand Function Test; JTT) and functional mobility (Timed Up and Go; TUG) following intervention. Positive values indicate behavioural improvement. Spearman correlation coefficients (rho), associated *p*-values, and 95% confidence intervals are displayed within each panel. No significant associations were observed between global electric field metrics and behavioural outcomes.

#### Grey Matter

No significant relationships were observed between global grey matter electric field strength (99.9th percentile magnE) and changes in JTT (rho = -0.105, p = 0.760) or TUG performance (rho = 0.205, p = 0.543; Figure 5). Similarly, stimulation focality, defined as the volume of grey matter exposed to electric field strengths equal to or exceeding 75% of the 99.9th percentile field, was not associated with changes in JTT performance (rho = -0.064, p = 0.860).

**Figure 5:**
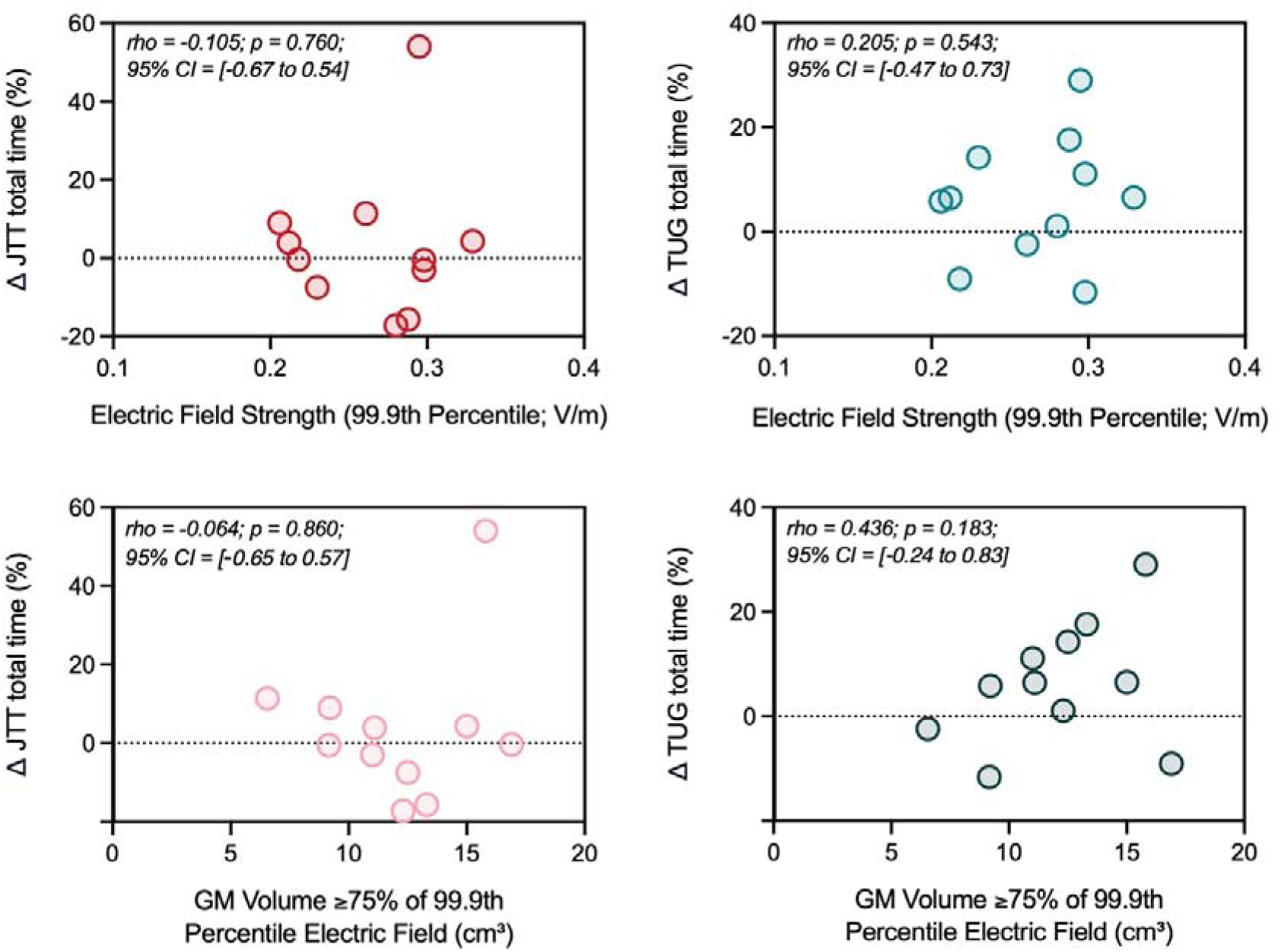
Scatterplots illustrating the relationships between global grey matter electric field metrics and behavioural outcomes within the active stimulation group. Upper panels show associations between the 99.9th percentile grey matter electric field magnitude (V/m) and changes in JTT and TUG performance. Lower panels show associations between stimulation focality and behavioural outcomes. Positive values indicate behavioural improvement. Spearman correlation coefficients (rho), associated p-values, and 95% confidence intervals are displayed within each panel. No significant associations were observed between global electric field metrics and behavioural outcomes.

A weak-to-moderate positive monotonic relationship was observed between a larger area of the grey matter receiving ≥75% electric field strength and change in TUG performance (rho = 0.436); however, this did not reach the statistical threshold for significance (p = 0.183).

## Discussion

The present study used individualised MRI-based finite element modelling to characterise electric field distributions generated by M1-targeted tDCS in young people with cerebral palsy. Consistent with the limited existing literature, subject-specific head models could be simulated successfully across a heterogeneous cohort with varying neuroanatomical pathologies [14, 25], including structural lesions. Three main findings emerged. First, estimated electric field strength differed significantly across motor regions, with PMd consistently receiving greater stimulation than both M1 and SMA despite the montage being designed to target M1. Second, electric field strength within a control region (V1) was substantially lower than within motor regions, supporting the relative anatomical specificity of the stimulation montage. Third, neither regional nor global electric field metrics were associated with improvements in upper- or lower-limb function following intervention. Collectively, these findings suggest that the actual distribution of current may differ from the intended cortical target and that inter-individual variability in modelled electric field strength alone does not explain variability in behavioural response in the present study.

Although the stimulation montage was designed to target M1, PMd consistently received the greatest electric field strength across participants. This finding highlights an important distinction between the intended cortical target and the possible distribution of current within the brain. Conventional tDCS montages employing large sponge electrodes in similar locations are known to produce relatively diffuse electric fields, with current distributed across multiple neighbouring cortical regions rather than being restricted to a single anatomical target [19, 38–40]. Consequently, some overlap in stimulation between adjacent motor regions would be expected and should not necessarily be interpreted as a phenomenon unique to CP. Indeed, diffuse current spread has been reported in modelling of adult stroke [41, 42].

Importantly, our modelling was successfully completed across a clinically heterogeneous cohort that included participants with differing CP aetiologies and motor phenotypes, including both unilateral and bilateral motor impairments. This extends previous paediatric CP modelling studies, which have largely focused on more homogeneous populations, while producing electric field magnitudes and focality measures comparable to those reported in children with perinatal stroke [25]. Together, these findings suggest that the observed predominance of PMd likely reflects both the characteristics of the stimulation montage and individual anatomy, rather than representing a phenomenon unique to CP.

One potential explanation relates to the electrode positioning employed in the present study. Unlike many upper-limb tDCS studies that centre the anode over C3/C4 [15], the electrode was positioned over C1/C2 to maximise coverage of both upper- and lower-limb motor representations. This more medial electrode position may have altered the spatial distribution of current within the motor network towards PMd. Nevertheless, electric field distributions are strongly affected by individual cortical geometry, including both gyri and sulci anatomy, tissue boundaries, and cerebrospinal fluid distribution [18, 20, 43].

Consequently, peak electric fields do not necessarily occur directly beneath the centre of the stimulating electrode. Previous computational modelling studies have similarly demonstrated discrepancies between intended and actual stimulation targets, with neighbouring cortical regions frequently receiving equivalent or greater stimulation than the target region [44–46]. The present findings extend these observations to a paediatric CP cohort and suggest that scalp-based methods of montage selection may overestimate the specificity of targeted stimulation.

Analysis did reveal significantly lower V1 electric fields (control region) supporting the relative anatomical specificity of the montage and suggesting that stimulation was preferentially concentrated within motor regions rather than distributed uniformly throughout the cortex. Importantly, while M1 remained relatively strongly stimulated, the consistently greater electric field strength within PMd raises the possibility that behavioural or neurophysiological effects attributed to M1 stimulation in the wider literature may instead reflect modulation of a broader motor network. This is particularly noteworthy given the strong functional connectivity between PMd and M1 and the recognised role of PMd in motor planning/preparation, processes that are central to rehabilitation interventions.

Despite inter-individual variability in modelled electric field strength, no significant relationships were observed between modelled electric field metrics and behavioural improvements following intervention. This was consistent across regional measures (M1, PMd, and the combined motor network ROI) and global measures of grey matter electric field strength and focality. These findings suggest that variability in electric field magnitude alone does not explain behavioural response to motor training within the present cohort. These findings are consistent with the broader tDCS literature, in which direct relationships between modelled electric field strength and behavioural improvement have been inconsistently demonstrated. While several studies have reported associations between individual electric field strength and neurophysiological measures such as functional connectivity [47, 48], evidence linking modelled electric fields to clinical motor outcomes remains limited and mixed.

Several explanations may account for this observation. First, behavioural responsiveness to tDCS is likely influenced by multiple factors beyond the anatomical impact on electric field strength, including baseline motor function, condition pathology and severity, lesion characteristics, plasticity, and the activities completed during the stimulation period. Consequently, the relationship between delivered electric field strength and behavioural outcome may be more complex than a simple dose-response association. Second, all participants received tDCS in combination with 7-10 sessions of upper- and lower-limb motor training. As motor training itself is known to induce meaningful functional improvements, any contribution of variability in electric field magnitude may have been relatively small compared to the effects of the behavioural intervention. It should be noted however, that no group effect of improvement in either JTT or TUG was observed between baseline and 1-week post-intervention in the original study [26]. It is possible that the intervention did not deliver large enough behavioural effects to detect any meaningful variation in response that could be related to stimulation-dose effects and therefore makes it difficult to untangle any intervention versus stimulation-dose effects in the present analyses. Third, the behavioural outcome measures employed in the present study may not have been sufficiently sensitive to detect subtle differences in tDCS response attributable to variation in the electric field delivered.

Importantly, these findings should not be interpreted as evidence that electric field strength is unrelated to treatment response. Rather, the present study may have been underpowered to detect such associations, particularly given that behavioural analyses were restricted to the active stimulation group (n = 11). Larger studies incorporating behavioural, neurophysiological, and imaging outcomes will be required to determine whether specific electric field characteristics are predictive of responsiveness to tDCS.

Inter-individual variability was also observed in global grey matter measures of electric field strength and stimulation focality. Despite this, neither electric field strength nor focality was significantly associated with changes in upper- or lower-limb function. Although a weak-to-moderate positive relationship was observed between a greater area of the grey matter receiving stronger electric fields and improvements in TUG performance - which may reflect the whole-body nature and thus required cortical engagement of an assessment like the TUG - this association did not reach statistical significance. Collectively, these findings suggest that global grey matter characteristics of current delivery may be insufficient to explain variability in behavioural response and reinforce the importance of considering precise stimulation targets, protocols, and timing when designing tDCS studies.

Several limitations of the present study should also be acknowledged. First, as a secondary analysis of data from an optional component of the original study, the cohort was relatively small (n = 19), and behavioural analyses were further restricted to participants receiving active anodal stimulation (n = 11), limiting statistical power to detect dose-response relationships. Second, only two participants with cortical lesions were identified, preventing meaningful investigation of the influence of lesion characteristics on electric field distribution. Finally, electric field modelling was performed retrospectively and therefore did not inform electrode placement or montage optimisation during the intervention. Individualised leadfield-based stimulation approaches [49] may improve targeting of specific cortical regions and could provide greater insight into the relationship between electric field characteristics and behavioural outcomes. Future studies incorporating larger cohorts, prospective electric field modelling, and personalised stimulation protocols are warranted to further investigate the determinants of tDCS responsiveness in young people with cerebral palsy.

In conclusion, individualised MRI-based electric field modelling demonstrated that a M1-targeted tDCS montage may result in significantly greater stimulation within PMd than either M1 or SMA in young people with cerebral palsy. Despite inter-individual variability in electric field strength and focality, neither region of interest nor global grey matter electric field metrics were significantly associated with behavioural improvements following intervention. These findings highlight the importance of subject-specific modelling when characterising the spatial distribution of current delivered by tDCS and suggest that conventional scalp-based targeting approaches may overestimate the specificity of stimulation to M1.

## Data Availability

All data produced in the present study are available upon reasonable request to the authors

## Abbreviations

(tDCS): Transcranial direct current stimulation;
(CP): Cerebral Palsy;
(M1): Primary Motor Cortex;
(PMd): Premotor Cortex;
(SMA): Supplementary Motor Area;
(ROI): Regions of Interest;
(MRI): Magnetic Resonance Imaging

## Rights Retention

This research is funded in whole, or in part, by the Wellcome Trust [222446/Z/21/Z, 203139/Z/16/Z and 203139/A/16/Z, 224430/Z/21/Z]. For the purpose of open access, the author has applied a CCBY public copyright license to any Author Accepted Manuscript version arising from this submission.

## CRediT roles

Conceptualisation: MW, BG, MKF

Methodology: MW, BG

Formal Analysis: MW

Data Curation: MW, BG, FM, MKF

Writing – original draft: MW

Writing – review and editing: MW, BG, FM, HJB, HD, MKF

Project administration: MW, BG, FM, MKF, HD

Funding: MKF, HJB, HD

